# Label-free virtual peritoneal lavage cytology via deep-learning-assisted single-color stimulated Raman scattering microscopy

**DOI:** 10.1101/2024.01.17.24301416

**Authors:** Tinghe Fang, Zhouqiao Wu, Xun Chen, Luxin Tan, Zhongwu Li, Jiafu Ji, Yubo Fan, Ziyu Li, Shuhua Yue

## Abstract

Clinical guidelines for gastric cancer treatment recommend intraoperative peritoneal lavage cytology to detect free cancer cells. Patients with positive cytology require neoadjuvant chemotherapy instead of instant resection and conversion to negative cytology results in improved survival. However, the accuracy of cytological diagnosis by pathologists or artificial intelligence is disturbed by manually-produced, unstandardized slides. In addition, the elaborate infrastructure makes cytology accessible to a limited number of medical institutes. Here, we developed CellGAN, a deep learning method that enables label-free virtual peritoneal lavage cytology by producing virtual hematoxylin-eosin-stained images with single-color stimulated Raman scattering microscopy. A structural similarity loss was introduced to overcome the challenge of existing unsupervised virtual pathology techniques unable to present cellular structures accurately. This method achieved a structural similarity of 0.820±0.041 and a nucleus area consistency of 0.698±0.102, indicating the staining fidelity outperforming the state-of-the-art method. Diagnosis using virtually stained cells reached 93.8% accuracy and substantial consistency with conventional staining. Single-cell detection and classification on virtual slides achieved a mean average precision of 0.924 and an area under the receiver operating characteristic curve of 0.906, respectively. Collectively, this method achieves standardized and accurate virtual peritoneal lavage cytology and holds great potential for clinical translation.

## Introduction

Gastric cancer (GC) is the third most common cause of cancer-related deaths^1^. According to clinical guidelines, GC staging should include laparoscopy-guided biopsy to detect peritoneal metastasis (PM) and intraoperative peritoneal lavage cytology to detect free intraperitoneal cancer cells. The disease is considered to be metastatic when either of them is positive. Patients with positive cytology require palliative neoadjuvant chemotherapy to eradicate free cancer cells instead of undergoing resection surgery immediately^2–4^. Then, conversion from positive to negative cytology is a significant predictor of better postoperative survival^5,6^. However, the application of cytology is impeded by problems caused by chemical slide processing. On the one hand, the conventional staining procedure has elaborate protocols and usually takes approximately 20 min. Although several modified versions of cytological staining (e.g., ultrafast Papanicolaou stain and Diff-Quik) reduce the time consumption to a few minutes, they can suffer from deficient diagnostic features or sensitivity to operations^7,8^. Moreover, they cannot overcome the inherent variation in image properties caused by inevitable differences in slide processing and scanning conditions. Such a lack of standardization significantly affects the universality of trained machine learning models in diagnosing whole-slide images. For example, variations in color style must be handled with color augmentation or normalization^9,10^. On the other hand, it is expensive to construct and maintain a certificated cytology laboratory with precision facilities and trained personnel^11^. Along with a number of cytology tests per year, the high consumption of hazardous reagents, such as xylene, also poses high economic costs in storage and disposal, as well as chemical hazards to pathologists^12,13^.

To avoid similar drawbacks of histology, including unstandardized image quality and elaborate infrastructure, several virtual pathology techniques have been developed. They are based on a variety of label-free microscopy techniques that provide contrasts among sample components, including autofluorescence imaging (AFI)^14–16^, photoacoustic microscopy (PAM)^17^, quantitative phase (QP) microscopy^18^, deep-ultraviolet (UV) microscopy^19^, and stimulated Raman scattering (SRS) microscopy^20–24^. Although they work well in virtual histology, most do not fit with the cellular scale for virtual cytology. The dependence of AFI on biomolecules such as FAD and NADH in biological samples results in only scattered signals in the cells^25^. PAM does not have a sufficient spatial resolution for cellular imaging^26^. QP microscopy lacks chemical specificity and thus cannot provide sharp intracellular contrast. Deep-UV microscopy causes photodamage to free cells and provides limited chemical information due to dependence on light absorption by a few biomolecules. In comparison, SRS has a higher potential for cellular imaging with subcellular spatial resolution and chemical-bond-level molecular specificity. In our previous work^27^, with SRS microscopy, we established a label-free PM detection method that matched well with the gold standard, laparoscopy-guided biopsy, and according to clinical guidelines, a corresponding SRS-based peritoneal lavage cytology with virtual staining is worth exploring. Currently, SRS microscopy can provide histological contrast in tissue sections as hematoxylin-eosin (H&E) stain does^28–30^. Its main drawback, equipment complexity, has been significantly mitigated by the development of fiber lasers and data acquisition schemes. Stimulated Raman histology (SRH), allowing virtual H&E staining, reduces the required Raman data for virtual pathology from hyperspectral SRS to two Raman shifts, that is, two-color SRS, and developed a fiber-laser-based imaging device facilitating surgical use^20–22^. A recent study further reduced the required data to images at one Raman shift, with which two-color images were predicted using deep learning (DL)^23^. In this case, a sophisticated dual-modal optical system combining picosecond and femtosecond lasers was required to construct the dataset. Nevertheless, as far as our knowledge goes, there has yet to be virtual pathology using single-color picosecond SRS images, so an unprecedentedly miniaturized fixed-wavelength laser cannot be used to facilitate clinical translation. As shown in previous studies^14,18,31,32^, virtual pathology can be achieved with single-channel label-free images using generative adversarial networks (GANs). A detailed comparison of the current imaging modalities for virtual pathology with staining is presented in **Table S1**.

In this study, we developed virtual stimulated Raman cytology (VSRC). It created virtual H&E-stained images of exfoliative cells in ascites by the developed staining model CellGAN, which was characterized by two points. On the one hand, single-color SRS was verified to be sufficient for virtual H&E staining. On the other hand, based on CycleGAN^33^, which incorporates adversarial loss for realistic staining and cycle consistency loss for the regularization of mapping reversibility, the model was trained without paired images. In particular, as an adaptation to imaging patterns of cytology from histology, structural similarity (SSIM) loss was introduced to CycleGAN for proper nucleocytoplasmic staining of free cells. The dataset of CellGAN included 7319 exfoliated cells from 32 ascites specimens. Diagnosis using virtually stained cells achieved an accuracy of 93.8% and substantial consistency with conventional chemical staining, as quantified by Cohen’s Kappa of 0.782. Two DL models were trained for downstream single-cell analysis based on virtually stained slides, achieving a mean average precision (mAP) of 0.924 in single-cell detection and an area under the receiver operating characteristic curve (AUC) of 0.906 in single-cell classification. Benefitting from the easy construction of an unpaired dataset and the CH_2_ vibration intrinsic to lipids common in all cells, our method holds great potential for application in other cytological scenarios.

## Results

### Workflow of CellGAN-assisted virtual stimulated Raman cytology (VSRC)

The workflow of CellGAN-assisted VSRC is shown in **Fig. 1**. 1) First, single-color SRS microscopy was first performed on the slides of exfoliated cells to avoid being affected by H&E staining. Virtual histology often requires large-scale image acquisition owing to the considerable heterogeneity of tumor tissues. In contrast, smaller coverage is required for cytology because free malignant cells can be captured more easily when mixed evenly with benign cells. As demonstrated in our previous study^27^, a 0.72 mm² SRS image costing only 1.5 min and containing ∼600 cells can be sufficient for the diagnosis of each patient. After SRS microscopy, the slides were stained with hematoxylin and eosin. 2) Second, the SRS and H&E-stained FOVs of each slide were stitched to form unregistered images for the training of CellGAN. Registered images were then obtained by cell-wise registration on a small number of unregistered images for validation and testing of CellGAN. A diagram of the unpaired and paired datasets is shown in **Fig. S1**. 3) Third, CellGAN, a DL model for virtual cytological staining, was trained using the CycleGAN framework. In addition to classical losses of CycleGAN, including cycle consistency loss and adversarial loss, the SSIM loss between images before and after translation by a generator was introduced to preserve the consistency of nucleocytoplasmic structures, which provide vital information for cytology. 4) Fourth, the staining performance of CellGAN was quantified using metrics and pathologist-assisted evaluation. Metrics reflecting overall fidelity and detailed nucleocytoplasmic consistency were calculated and compared with the UTOM^15^, the baseline model, on the test dataset. UTOM is an unsupervised image translation framework based on CycleGAN and has been used to create virtual H&E-stained tissue sections in previous studies^16,19^. Assisted by pathologists, the diagnostic value of virtually stained cells was evaluated according to whether they contained consistent information relative to cells stained using conventional chemical processing. 5) Fifth, DL models for single-cell analysis after virtual staining were trained to detect all cells in a virtually stained slide image and then classify them into benign and malignant groups. The detection and classification results were compared with the manual annotations of the corresponding chemically-stained slides.

**Fig. 1.**
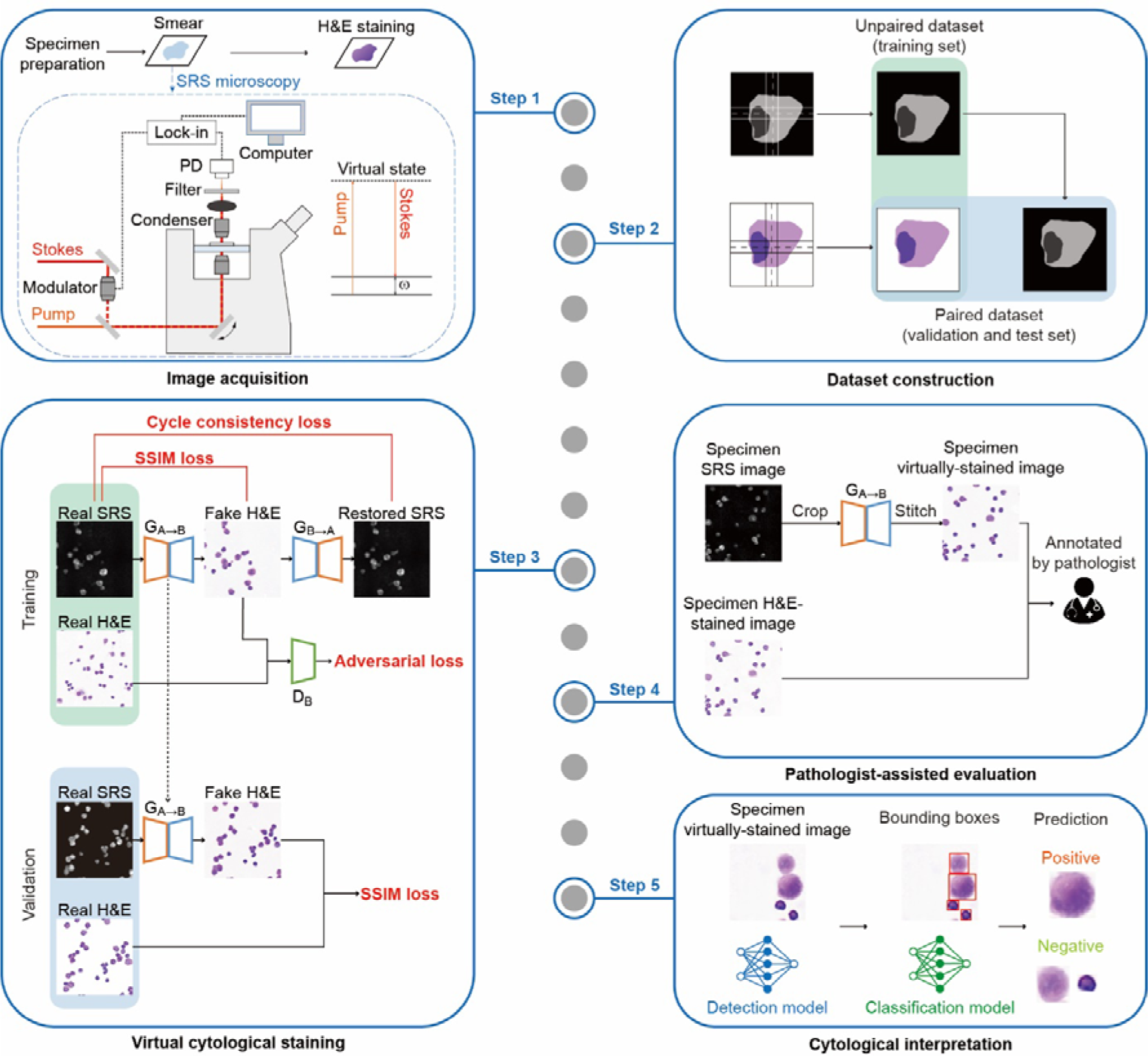
Workflow of CellGAN-assisted virtual stimulated Raman cytology (VSRC). Step 1, specimen preparation, SRS imaging, and H&E staining. Step 2, stitching and registration of SRS/H&E-stained images to construct unpaired and paired datasets. Step 3, training of CellGAN based on the CycleGAN. Step 4, pathologist-assisted evaluation of CellGAN-assisted VSRC. Step 5, training of subsequent DL models for single-cell analysis, including single-cell detection and classification.

### Dataset construction: Single-color SRS cytological imaging, stitching, and registration

Single-color SRS microscopy was performed by tuning the laser wavelengths to match the Raman band of the CH_2_ symmetric stretching vibration at 2850 cm^-1^, which created a strong contrast between lipid-rich cellular structures (e.g., cell membranes) and lipid-poor nuclei. As shown in **Fig. S2A**, FOVs belonging to the same slide were stitched together with a 30% overlap specified when these FOVs were captured. Seams between every pair of adjacent FOVs were then smoothed by linear blending to mitigate lattice artifacts which were mainly caused by uneven illumination. Smoothing inhibited sudden pixel intensity changes near the seams considerably, as seen in the stitching result (**Fig. 2A**) and its intensity profile (**Fig. 2B**). Quantitatively, Sobel gradients that quantify the sharpness of intensity changes were found to significantly decrease on seams after smoothing (**Fig. S2B**).

**Fig. 2.**
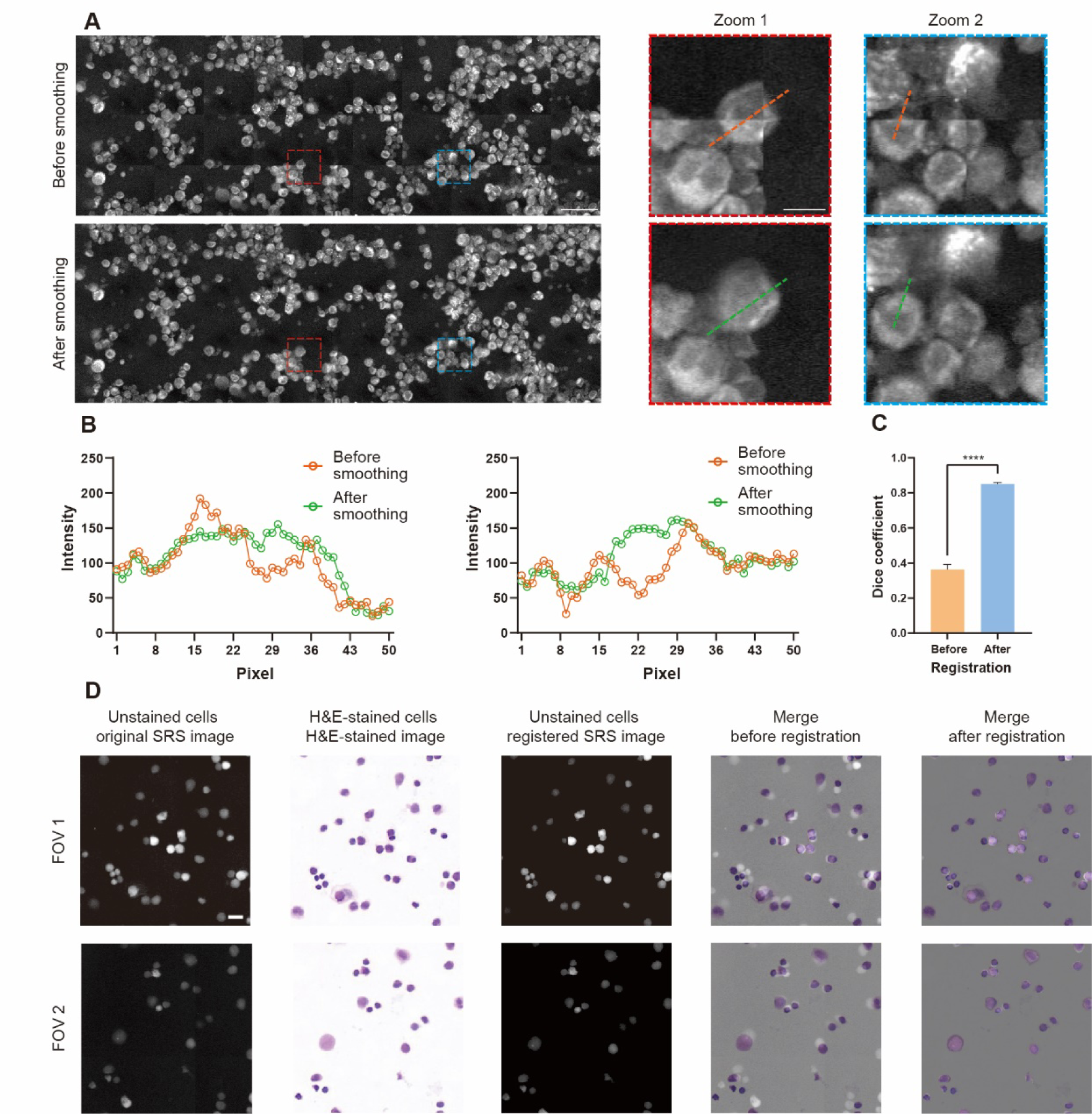
Image stitching and registration in dataset construction. **A**, Mosaic image before and after seam smoothing. Overall-image scale bars, 50 μm. Close-up scale bars, 10 μm. **B**, Intensity profiles along orange and green dashed lines in Zoom 1 (red dashed border) and Zoom 2 (blue dashed border) in **A**. **C**, Dice coefficients between pairs of binarized patches (n=100) before (mean±SD=0.362±0.030) and after registration (mean±SD=0.850±0.009). **D**, SRS/H&E-stained patches before and after registration. Scale bars, 10 μm.

SRS and H&E-stained stitched images were obtained from 32 slides. Images from 24 slides were randomly cropped into ∼5000 SRS patches and the same number of H&E-stained patches. They constituted the training dataset of CellGAN, which was unpaired because of the inevitable cell shift or loss during chemical staining. Images from the other eight slides were used to generate the validation and test datasets of CellGAN, which were paired after being processed by a registration algorithm. As shown in **Fig. S2C**, aspired by the algorithm developed previously^14^, our algorithm was composed of rough patch matching and iterative registration that included pyramidal partitioning and block-wise alignment. In addition, each block was adjusted to avoid cell truncation, as described in the Methods section. After iterative registration, the pairwise Dice coefficients of the patch binary masks significantly increased compared with those of the roughly matched masks (**Fig. 2C**). The representative patch pairs before and after iterative registration are shown in **Fig. 2D**. The representative registration process is shown in **Fig. S2D**. A total of ∼1200 registered patch pairs were generated and equally distributed into the validation and test datasets of CellGAN.

### CellGAN: Development of deep learning model to assist label-free virtual peritoneal lavage cytology

After training, the performance of CellGAN was preliminarily observed within the unpaired dataset, which did not involve data leakage due to the unsupervised quality. A representative pair of SRS and H&E-stained images of the same slide in the unpaired dataset is shown in **Fig. 3A** and **Fig. 3B**, respectively. Since the content-preserving loss in UTOM roughly divides the image into two parts, regions with and without salient features, to some degree, it cannot identify whether the nucleocytoplasmic structures are properly stained when the training is unsupervised. As a result, UTOM was found to frequently stain the nucleus and cytoplasm in reverse (**Fig. 3C**), thus producing misleading outputs. To explore a solution to this problem, we conducted an experiment where simulative SRS and H&E-stained cells were drawn with empirical colors of cell structures, as shown in **Fig. S3A**. According to its results, we hypothesized that reversely stained images were less structurally similar to unstained SRS images than properly stained images, as quantified by SSIM. This critical finding suggests that SSIM loss may help to avoid the reverse staining problem. Heuristically, we tested our hypothesis by replacing the content-preserving loss in the UTOM with the SSIM loss. It was inspiring that CellGAN produced virtual cytological images that were highly similar to the ground truth (GT) (**Fig. 3D**). With respect to the mapping of nucleocytoplasmic structures, the performance of CellGAN was better than that of UTOM (**Fig. 4A**).

**Fig. 3.**
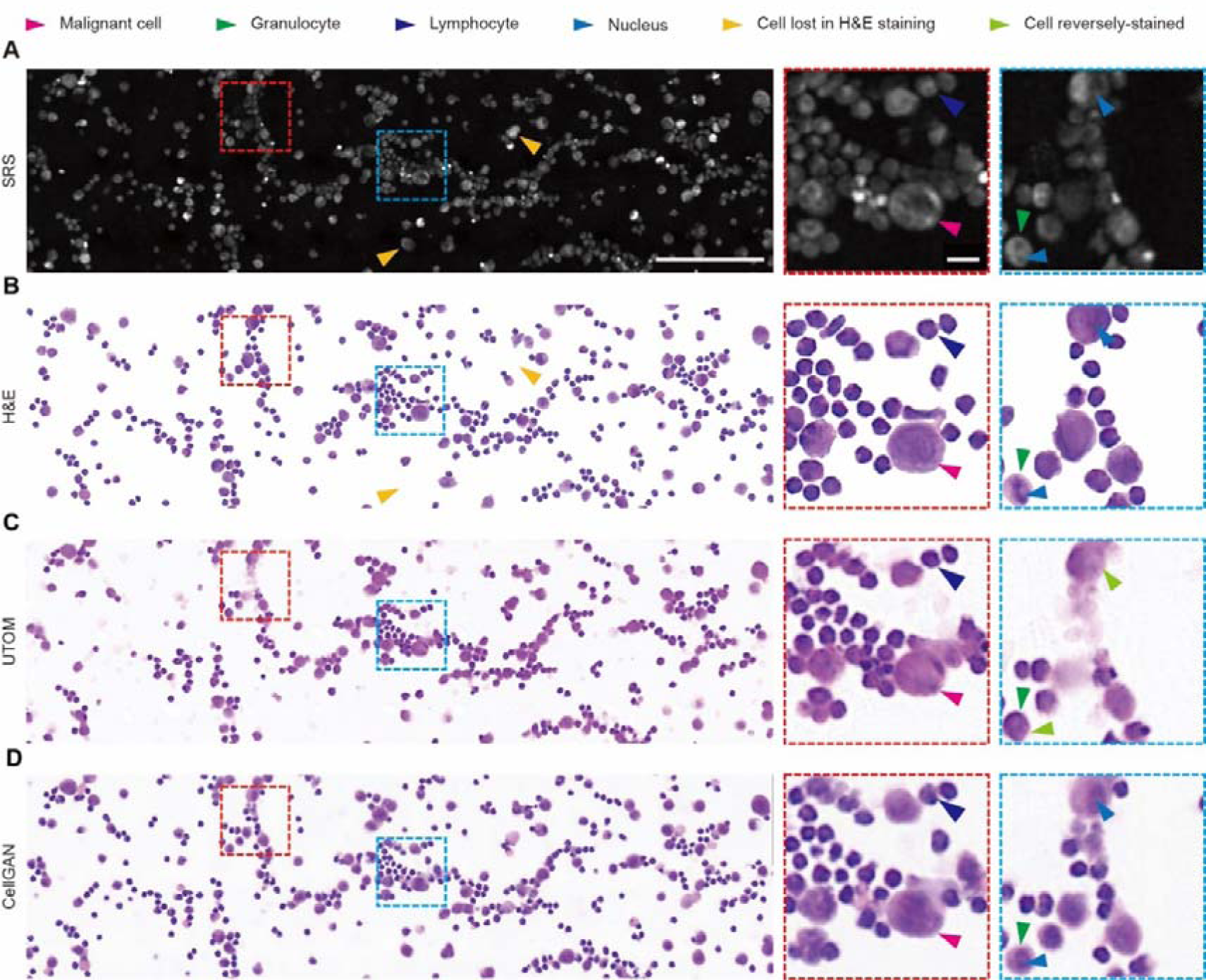
Virtual cytological staining result matching the H&E stain for ascites fluid slides. **A**, SRS image of an unprocessed ascites fluid slide. **B**, The same slide stained with H&E stain after registration with **A**. **C**, The same slide virtually stained by UTOM. **D**, The same slide virtually stained by CellGAN. Overall-image scale bars, 100 μm. Close-up scale bars, 10 μm.

**Fig. 4.**
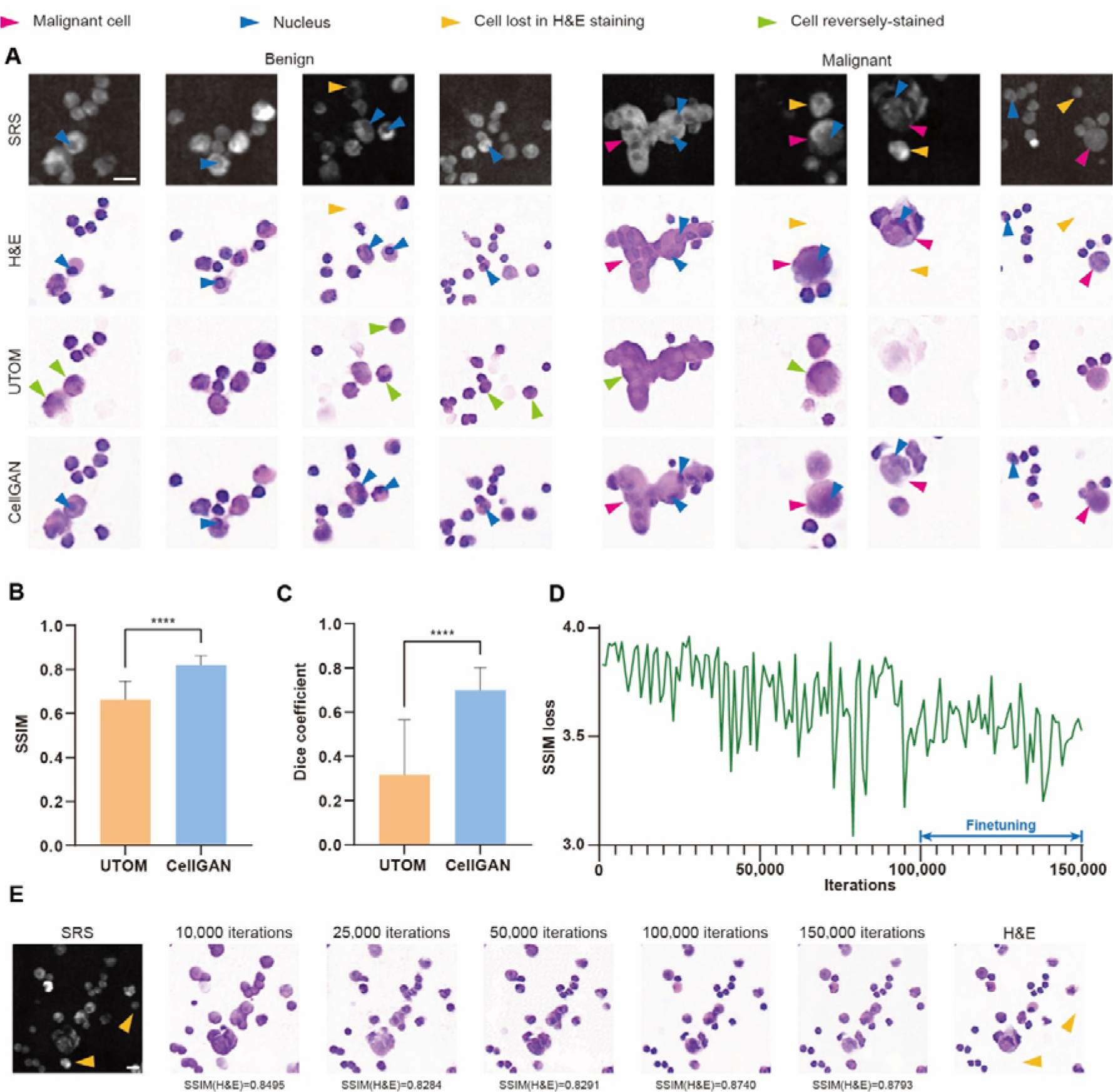
Qualitative and quantitative analysis of virtually stained cells. **A,** Exfoliated cells that are unstained, H&E-stained (GT), virtually stained by UTOM and CellGAN. **B**, SSIMs between virtually stained images and the GT (n=618) produced by UTOM (mean±SD=0.662±0.085) and CellGAN (mean±SD=0.820±0.041). **C**, Dice coefficients of nucleus regions between virtually stained images and the GT (n=618) produced by UTOM (mean±SD=0.315±0.249) and CellGAN (mean±SD=0.698±0.102). **D**, The SSIM loss against the number of iterations in the training process. **E**, Evolution of cells virtually stained by CellGAN as the number of iterations increased. Scale bars, 10 μm.

Next, we quantitatively evaluated the performance of CellGAN in terms of staining style, cell morphology, and nucleocytoplasmic structures on the test dataset. These features provide vital information for cytological diagnosis. SSIMs between the model outputs and the GT reflected general similarity in staining style and cell morphology. The values were larger for CellGAN (0.820±0.041) than for UTOM (0.662±0.085) (**Fig. 4B**). As a more critical and fine-grained metric, Dice coefficients reflected nucleocytoplasmic consistency by comparing the nuclear region of each cell in the model outputs and the GT, which was extracted with an empirical threshold. These values were remarkably larger for CellGAN (0.698±0.102) than for UTOM (0.315±0.249) (**Fig. 4C**). These results suggest that CellGAN can achieve virtual cytological staining with higher fidelity at different scales. We also compared our results with SRH (**Fig. S4**), which were calculated using single- and two-color SRS, respectively. Both matched well with H&E-stained images in terms of cellular structures and cell types, including granulocytes and lymphocytes. Therefore, our single-color picosecond SRS scheme can simplify data acquisition significantly with similar virtual staining quality.

Moreover, we analyzed how SSIM contributed to proper nucleocytoplasmic mapping. First, SSIMs between model outputs and unstained SRS images were compared (**Fig. S3B**). It was found that although properly stained and reversely stained images had similar color styles, properly stained images were significantly more similar to inputs, as quantified by the SSIM. This finding conforms to our hypothesis and implies that the excellent performance of CellGAN could be associated with the SSIM loss. We then tracked the evolution of the SSIM loss (**Fig. 4D**) and CellGAN outputs (**Fig. 4E**) during the training process. At first, proper nucleocytoplasmic mapping was learned rapidly, as indicated by the CellGAN output after ∼10 thousand iterations. However, at that moment, all cells appeared to be stained homogeneously, meaning that cells of different types could hardly be distinguished. As training continued, differences among different cell types became more prominent, and nucleocytoplasmic contrast was further enhanced. Therefore, we gained more insights into the training process of CellGAN with the guidance of the SSIM loss.

### Pathologist-assisted evaluation of CellGAN-assisted VSRC

After evaluating the staining fidelity of CellGAN-assisted VSRC, we demonstrated the diagnostic value of virtually stained cells in the cytological workflow. For the dataset containing 32 slides, 7319 exfoliated cells were virtually stained using CellGAN and then manually annotated by two pathologists. In addition, H&E-stained images produced by conventional chemical staining in the whole dataset were annotated separately from the virtually stained ones. Therefore, each cell, except those lost in chemical staining, had two annotations, whose consistency reflected whether virtually stained slides conveyed diagnostically relevant information as chemically-stained slides^34^.

The confusion matrices of cell-level and slide-level diagnoses are shown in **Table 1**. The cell-level diagnosis, where annotations of each cell were considered, achieved 99.4% true negative, 99.7% true positive, and 99.4% accuracy. The definitions of these three metrics are described in the Materials and Methods section. Cohen’s Kappa calculated for each slide, which reflected consistency between diagnosis based on virtual and chemical staining, was 0.782±0.284, indicating substantial consistency. These results indicate the applicability of CellGAN-assisted VSRC for cytological diagnosis.

**Table 1.** Cell-level and slide-level diagnoses by pathologists based on virtual and chemical staining.

|  |  | Chemical staining |  |  |  |
| --- | --- | --- | --- | --- | --- |
|  |  | PM Negative slides |  | PM Positive slides |  |
| Virtual staining | PM Negative slides | 20 |  | 0 |  |
|  | PM Positive slides | 2 |  | 10 |  |
|  |  | Benign cells | Malignant cells | Benign cells | Malignant cells |
|  | Benign cells | 5305 | 0 | 1585 | 1 |
|  | Malignant cells | 9 | 0 | 35 | 384 |

Moreover, the statistics of slide-level diagnosis were also recorded because the accurate diagnosis of a slide is closer to clinical requirements than that of an individual cell. The slide-level diagnosis, in which only slide annotations inferred from cellular ones were considered, achieved 100% true positive, 90.9% true negative, and 93.8% accuracy. The Cohen’s Kappa was 0.862, indicating an almost perfect consistency. Therefore, we demonstrated that CellGAN-assisted VSRC could represent cytological features that are recognizable by pathologists for accurate diagnosis.

### Deep learning-based single-cell analysis in CellGAN-assisted VSRC

We further trained a cascade of DL models for single-cell analysis to test the feasibility of downstream cytological interpretation based on the CellGAN-assisted VSRC. In this task, each cell was detected as a bounding box and then classified into benign or malignant groups.

In the first step, we trained a YOLOv5-based model to detect individual cells. As shown in **Fig. 5A-C**, the detected bounding boxes conformed well to manual annotations. Quantitatively, the intersection over union (IoU), which reflects the extent of overlap between two regions, was calculated from each bounding box and its corresponding annotation (**Fig. 5D**). Most IoU values were greater than 0.7. Zero values corresponded to wrongly-detected faint shadows resulting from out-of-focus cells in the unstained SRS image. Quantitatively, the detection model achieved a mean average precision (mAP) of 0.924, with the precision-recall curve shown in **Fig. 5E**. In the second step, we trained a ResNet50-based model to classify each cell cut from detected bounding boxes. Before cells were fed into the classification model, wrongly-detected cells could be filtered out with an empirically-set intensity threshold due to their whitish color. Other cells were then classified, as shown in **Fig. 5F-I**. The classification model achieved an accuracy of 96.7% and an AUC of 0.906 (**Fig. 5J**). These results suggest that CellGAN-assisted VSRC could fit well in the digital cytological workflow.

**Fig. 5.**
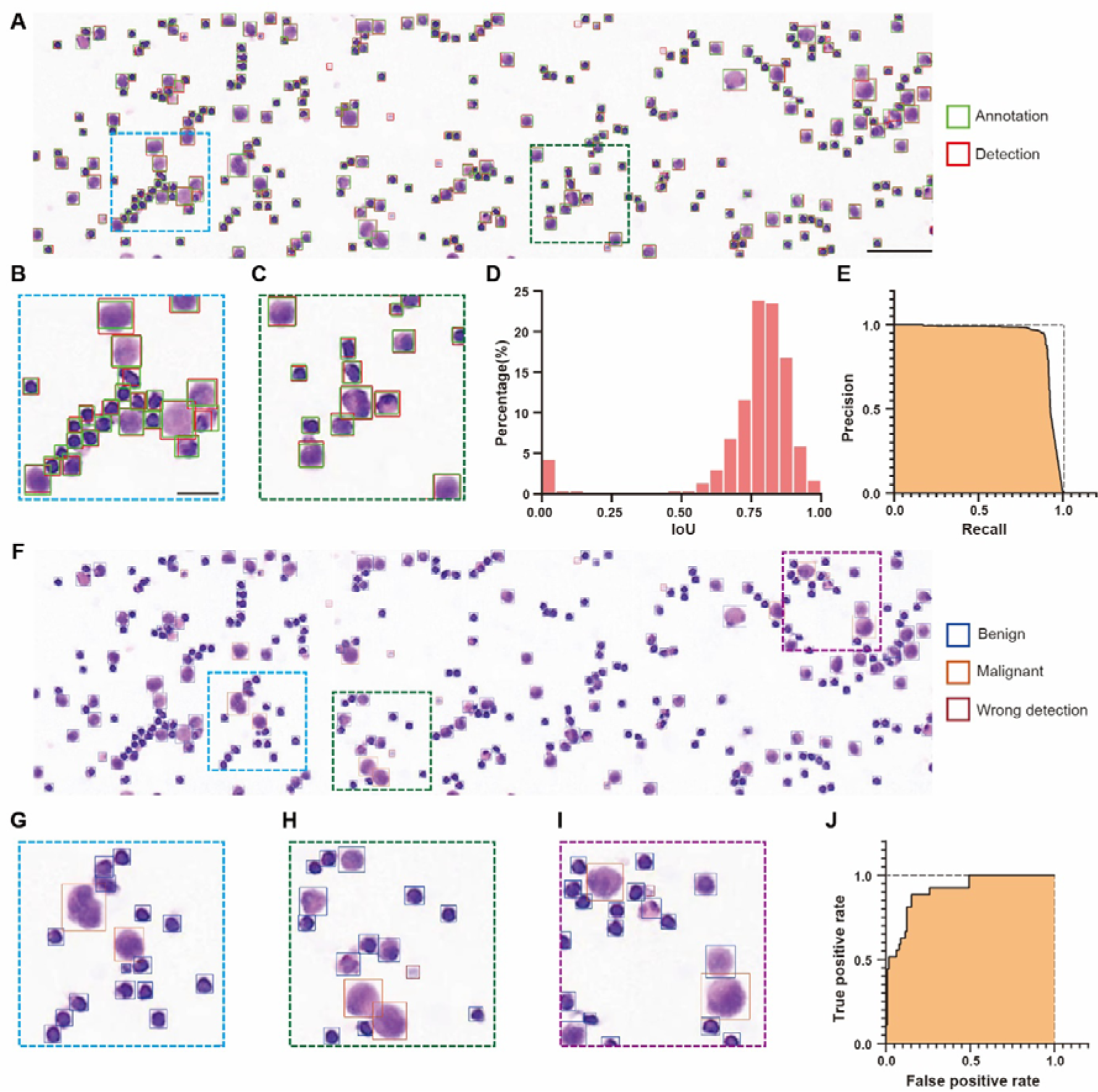
Deep learning-based single-cell analysis in CellGAN-assisted VSRC. **A**, Cell detection results and manual annotations of a virtually stained slide. **B, C**, Close-ups of **A**. **D**, IoU between detected bounding boxes and matched annotations (n=311). **E**, Precision-recall curve of cell detection. **F**, Classification results of detected cells in the same slide. **G-I**, Close-ups of **F**. **J**, ROC curve of cell classification. Overall-image scale bars, 200 μm. Close-up scale bars, 50 μm.

## Discussion

In this study, we developed CellGAN enabling VSRC, a label-free virtual peritoneal lavage cytology. Trained on an unpaired dataset, CellGAN realizes virtual cytological staining by learning the mapping between single-color SRS and H&E-stained images. The applicability of the virtually stained cells was evaluated through diagnosis by pathologists and additional DL models for downstream single-cell analysis. The CellGAN-assisted VSRC realized a standardized and automatic digital cytological workflow. As discussed below, our study may open up new avenues for cytology.

First, the proposed method realizes virtual cytology without wavelength tuning. As established in previous studies, imaging at multiple Raman shifts is frequently required to decouple different chemical components for further analysis^29,35^. However, along with rich spectroscopic information, multi-color SRS also causes higher complexity of laser sources and lower environmental stability. Therefore, to a certain degree, the multi-color scheme impedes the clinical translation of SRS-based virtual pathology. In comparison, although single-color SRS microscopy contains less information, DL can be used to obtain the latent features for virtual staining. Therefore, our method can be implemented with a device that is as simple as a dual-fixed-wavelength picosecond pulse laser. Currently, compact fiber lasers have been developed for CRS microscopy^36–38^, making it promising to perform our method with an integrated, miniaturized, and portable imaging device.

Second, the cytological dataset used to implement our method is highly accessible. Benefitting from the unsupervised framework CycleGAN, the demand for data included in the dataset was less strict, meaning that images could be obtained at different locations on different slides using different equipment. Unsupervised learning is especially meaningful for cytology because the discreteness and deformability of cells elevate the cost of high-quality ground truth images. When using unsupervised learning, the image collection should be sufficiently representative to reflect the statistical features of both sides of the mapping. For example, overrepresentation or underrepresentation of one or several features (e.g., cell types) may cause biased image translation^39^; differences in the scales of cytological images can also mislead the model.

Third, we heuristically introduced the SSIM loss to CycleGAN and analyzed its contribution to the proper staining of cellular structures. Sometimes, CycleGAN requires losses that are more powerful than its classical cycle consistency loss to preserve image contents after mapping. Therefore, additional losses are introduced, such as the binary mask loss^15^ used by UTOM or perceptual loss^40^. The binary mask loss quantifies the discrepancy between the binary masks of salient features before and after image translation performed by a generator. It works well for virtual histological staining. However, when applied to cells, the nucleocytoplasmic information is neglected. Consequently, it is very likely to cause the reverse staining problem. In CellGAN, another form of content-preserving loss, the SSIM loss, was introduced. SSIM is related to the image texture, and thus fine-grained features can be handled better, especially for correcting nucleocytoplasmic mapping. This guarantees proper representation of cellular structures, which provide vital information for cytological diagnosis. Such a countermeasure reflects one of the challenges unique to virtual cytological staining compared to histology.

Tissue sections are characterized by apparent intrinsic organization and continuity, while cytological slides have different types of cells and scattered spatial distribution. For histology, continuous patterns of cell arrangement provide important candidate features for a virtual staining model, whereas cytology is very different. In this case, the challenging identification of intercellular variability and intracellular structures limits the performance of the existing virtual staining methods for histology when directly applied to cytology.

Fourth, our method is promising in combination with high-throughput techniques, extended cytological databases, and more ingenious configurations of DL models. On the one hand, our method’s upstream image acquisition can be improved with techniques like microfluidics and flow cytometry^41,42^. Coherent Raman imaging flow cytometry has achieved a throughput of thousands of events per second^43^. With moderate demand for imaging devices, our method can be easily combined with it for higher throughput. On the other hand, the ability of our model to process cells could be further improved. Although CellGAN achieved precise nucleocytoplasmic mapping, it could not recognize and process each cell individually explicitly, which may limit its performance by producing artifacts. For example, clustered cells may be misidentified as large malignant cells, and some indistinct cells may be neglected (**Fig. S5**). These can also be improved using cell-dispersion techniques and refined focusing.

In conclusion, our method holds great promise for developing into generalizable cytology with high-level standardization, accuracy, and automation. Because of the ubiquitous target Raman-active mode, that is, CH_2_ symmetric stretching, which reflects lipid distribution instead of cell-specific labels, our method can be extended to other cytological scenarios. Transfer learning may reduce the training costs in scenarios involving slides with similar cellular features.

## Methods

### Ascites specimen preparation

Peritoneal lavage fluids were collected from 11 patients (seven with negative cytology and four with positive cytology) diagnosed with locally advanced gastric cancer at the Peking University Cancer Hospital with informed consent. For each patient, 100 mL of fluid was collected. This study was approved by the Institutional Review Boards of Peking University Cancer Hospital and Beihang University.

After collection, ascites specimens were centrifuged at 2000 rpm for three minutes. They were then treated with red blood cell lysis buffer for five minutes and rinsed with phosphate-buffered saline. The specimen (∼10 μL) was dropped on a glass slide and evenly smeared over an area of ∼1 cm^2^ to facilitate subsequent H&E staining. After air-drying, the slide was used for SRS microscopy and stored at −80°C as a backup.

### Image acquisition and preprocessing

SRS was performed with a homebuilt multimodal nonlinear optical microscope. Pump and Stokes beams were outputted by a picosecond pulsed laser (picoEmerald^TM^ S, Applied Physics & Electronics) with an 80 MHz repetition rate and two picosecond pulse duration. The wavelength of the pump and Stokes beam was respectively fixed at 796.8nm and 1031nm to match the Raman band of CH_2_ symmetric stretching vibration at 2850 cm^-1^, which provided a stronger nucleocytoplasmic contrast than the Raman band of CH_3_ symmetric stretching vibration at 2930 cm^-1^. The Stokes beam was modulated at ∼20 MHz by an electronic optic modulator. The collinear pump and Stokes beams were coupled to a two-dimensional scanning galvanometer (GVS012-2D, Thorlab) and imported into an inverted microscope (IX73, Olympus). They were focused onto the specimen with a 60X water-immersion objective (LUMPlanFL N, 1.0 numerical aperture, Olympus) and then collected by another 60X water-immersion objective (LUMPlanFL N, 1.0 numerical aperture, Olympus). A short-pass filter (ET980SP, Chroma) was used to filter out the Stokes beam before a silicon photodiode (S3994-01, Hamamatsu) detector with 48 DC reversed bias voltage to receive the SRS signal, which was then extracted by a lock-in amplifier (HF2LI, Zurich Instruments). The analog output was fed into a data acquisition card (PCIE-6363, National Instruments), which inputted the signal to a computer. Finally, it was saved as a 400×400 image on the LabVIEW 2018 platform. The pixel dwell time was 10 μs. The FOV of each image was 100×100 μm.

Image stitching combined FOVs belonging to the same slide into a mosaic image. In the specified overlap, pixels belonging to two FOVs were linearly blended as

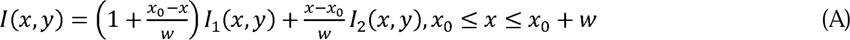

where *w* denotes the overlap width; [*x_0_*, *x_0_+w*] denotes the range of the overlap coordinates along its width; *I_1_* and *I_2_* denote the adjacent FOVs to be combined. The FOVs belonging to each row were combined, and then all rows were combined along the other axis into the stitching result.

In addition, the linearly blended area can be narrower than the specified overlap width to maintain the original intensity as much as possible. Then, either of the two adjacent FOVs determines the intensity of the remaining area. After stitching, the scales of the SRS and H&E-stained images should be aligned because of the difference in pixel resolution. After stitching, the data was cleaned by checking imaging quality with visual judgment. Then, other preprocessing included converting 32-bit to 8-bit RGB, contrast enhancement, and Gaussian filtering of stitched SRS images. The training dataset can then be constructed with randomly cropped SRS and H&E-stained mosaic images.

Image registration pyramidally aligned each cell in a mosaic image. Firstly, for a randomly selected patch in the stitched SRS image, a patch in the H&E-stained image of the same slide was roughly matched by the maxima of normalized cross correlation (NCC), a similarity measure commonly used for image matching. To note, before matching, the H&E-stained image should be converted into grayscale, and then its intensity was reversed because the background colors of SRS and H&E-stained images were opposite. Then, two matched patches were binarized to masks by an adaptive threshold (SRS) and a constant threshold (H&E) to separate each patch into foreground and background. Pixel intensity in the background of original patches was assigned 0 (SRS) and 255 (H&E) for all RGB channels.

Second, in each pair of binary masks, the SRS patch was divided into N² blocks (e.g., N=3). For each binary block B_i_ (i=1, 2,…, N^2^), its contents must be corrected in two steps:1) If there were truncated cells, the edge of the block where these truncated cells were located should be extended until they were included entirely. During the extension, if pixels belonging to new cells were included in the block, they should be removed by assigning zero values. 2) After the first step, if there were cells that had been processed in past blocks (B_1_, B_2_,…, B_i-1_), these cells should be removed. After block correction, the maxima of the NCC determined the optimized shift of the block along the two axes. Finally, after shifting all blocks, N was replaced by N+1, and the iteration described above was performed again on the latest result until the increment in accuracy was less than a threshold. At the end of each iteration, the shifting operation on the two binary masks was duplicated on the two patches to perform the registration.

Registration accuracy was quantified using the Dice coefficient between pairs of binary masks. Some cells may deviate from the correct location temporarily because they were distributed into a block with larger cells and thus neglected by optimization of NCC, but they gained attention later when N was large enough. After selecting a specified number of SRS patches and obtaining their corresponding registered H&E-stained patches, the validation and test datasets were constructed.

The Dice coefficient was defined as

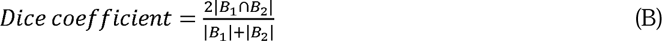

where *B_1_* and *B_2_* are binarized masks of two patches and |·| is the foreground area in a binary mask. The performance of the algorithm decreases as the cells deviate further and more chaotically. Slides with subtle cell shifts and nice registration after their patches were fed into the algorithm were chosen to generate validation and test datasets.

Images in the entire dataset had 256×256 pixels in an 8-bit RGB format to make generators and discriminators have the same architecture, respectively, as described below.

### Architecture and training of CellGAN

CellGAN was developed based on CycleGAN, which is composed of two generators and two discriminators. In CycleGAN, two image domains (e.g., SRS and H&E) represent two statistical distributions of image features, that is, two image styles. In our configuration, one generator learns the mapping from SRS to H&E, and the other learns from H&E to SRS. The discriminator should identify whether an input image is synthesized by its corresponding generator. Owing to the lack of paired data in the training dataset, losses based on pixel-wise differences cannot be used. The cycle consistency loss is introduced as a constraint that complements adversarial losses. It is defined as the difference between the input and reconstructed input obtained after the input is mapped into another domain and then mapped back.

In CellGAN, the generator is a residual network composed of an encoder, residual blocks, and a decoder, and the discriminator is 70×70 PatchGAN^44^. Their architectures are shown in **Fig. S6**. The kernel size was seven in the generators’ first and last convolutional layers and three in others. The models were built using PyTorch. Parameters were initialized randomly and optimized using Adam with a 1e-4 initial learning rate and beta values of 0.5 and 0.999. After ∼100 thousand iterations (∼20 epochs), the learning rate decayed to 1e-5 to fine-tune the parameters in another ∼50 thousand iterations (∼10 epochs). A NVIDIA GeForce RTX 3070 was used to train the model.

The losses of generators and discriminators were defined as (with empirically set parameters γ=10 and ρ=2)

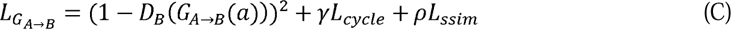

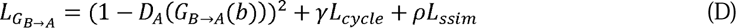

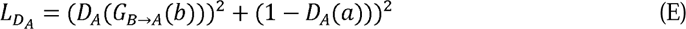

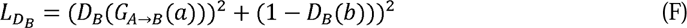

where *D_X_*denotes the discriminator identifying whether an image is synthesized by *G_Y_*_→*X*_ or from reality; *G_Y_*_→*X*_ denotes the generator converting an image from image domain *Y* into another image domain *X*; *a* and *b* respectively denote an SRS image and an H&E-stained image in the training dataset; *A* and *B* respectively denote the image domains of SRS and H&E.

The cycle-consistent loss and the SSIM loss were defined as

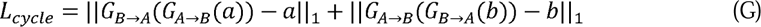

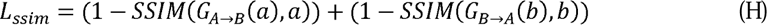

where ||·||_1_ denotes L1 loss and SSIM denotes structural similarity between two images. The SSIM was defined as

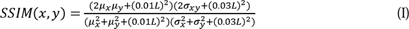

where *x* and *y* denote the convolution of the original images and a Gaussian kernel; μ_x_ denotes the average of *x*; μ*_y_* denotes the average of *y*; σ*_x_* denotes the standard deviation of *x*; σ*_y_* denotes the standard deviation of *y*; σ*_xy_* denotes the covariance of *x* and *y*; *L* denotes the range of pixel intensity (*L*=255 when images are in 8-bit RGB format).

### Pathologist-assisted evaluation

Produced by virtual and chemical staining, slide images were annotated by pathologists, and each cell was classified into benign or malignant groups. Because virtually stained images were acquired from unregistered SRS images, a cell may be in different locations on virtually stained and H&E-stained slides. The cellular correspondence between the cells was identified by visual judgment. As a two-dimensional binary classification, cellular annotations in the two types of stained slide images were recorded in a confusion matrix.

True positive rate, true negative rate, and accuracy were defined as

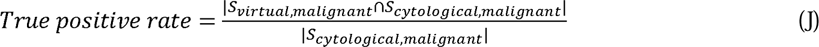

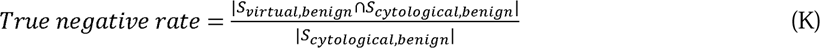

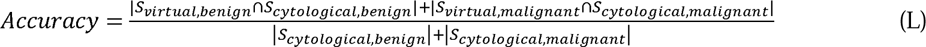

where *S* denotes the set of cells or slides with a certain annotation in virtual or chemical staining, as shown in the confusion matrices, and |*S*| denotes the number of elements in set *S*. These metrics are similar to the sensitivity, specificity, and accuracy for the evaluation of a binary classification model.

To compactly quantify the consistency between annotations in two types of stained slide images, Cohen’s Kappa was calculated using the confusion matrix as

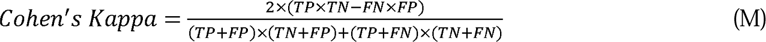

where TP, TN, FP, and FN denote the true positive, true negative, false positive, and false negative, respectively. For cell-level diagnosis, each slide corresponded to a Cohen’s Kappa value. For slide-level diagnosis, all slides corresponded to a Cohen’s Kappa value.

### Single-cell analysis with deep learning models

A YOLOv5-based model was trained with the pre-trained checkpoint YOLOv5s. Each virtually stained slide image was equally divided into three parts along its width. They were resized to 640 pixels in width and then padded into a square. The other parameters were the default values in the original code of YOLOv5. A ResNet50-based model was trained with pre-trained weights on ImageNet. The output size of the fully connected layer was modified to two for binary classification. The cross-entropy loss was employed to train the model. The optimizer was AdamW with an initial learning rate of 1e-4, which decayed exponentially with a gamma of 0.99. In the evaluation of cell detection, predicted bounding boxes were filtered with a 0.9 threshold of confidence to remove redundant bounding boxes that corresponded to the same cell. Each predicted bounding box matched the annotated bounding box that minimized the distance between their centers. The IoU between two bounding boxes is defined as

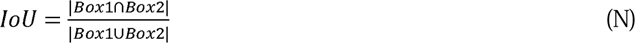

where |·| denotes the area of a region.

## Data Availability

All data produced in the present study are available upon reasonable request to the authors

## Acknowledgements

This work was supported by National Natural Science Foundation of China (No. 62027824, No. U20A20390, No. 11827803, No. 91959120, and No. 62205010), Natural Science Foundation of Beijing Municipality (No. 7224367 and No. L223018), Fundamental Research Funds for Central Universities (No. YWF-22-L-547 and No. YWF-22-L-1265), Higher Education Discipline Innovation Project (No. B13003), Clinical Medicine Plus X-Young Scholars Project of Peking University, and Science Foundation of Peking University Cancer Hospital.

## Author Contributions

**T. Fang:** Conceptualization, data curation, software, formal analysis, investigation, visualization, methodology, writing-original draft; **Z. Wu:** Resources, funding acquisition, methodology; **X. Chen:** Funding acquisition, investigation, methodology, writing-original draft; **L. Tan:** Resources, formal analysis; **Zhongwu Li:** Resources, formal analysis; **J. Ji:** Conceptualization, resources; **Y. Fan:** Conceptualization, supervision, funding acquisition, writing-review and editing; **Ziyu Li:** Conceptualization, supervision, writing-review and editing; **S. Yue:** Conceptualization, supervision, funding acquisition, investigation, methodology, writing-original draft, writing-review and editing.

