## Supplemental Table 1 and Figure 1-6 for "Label-free virtual peritoneal lavage cytology via deep-learning-assisted single-color stimulated Raman scattering microscopy"

**Table S1. Comparison of current label-free imaging modalities used for virtual pathology**

| Imaging modality | Source of contrast in samples | Advantages & Disadvantages |
| --- | --- | --- |
| Autofluorescence imaging | Endogenous fluorophores (e.g., collagen, elastin, NAD(P)H, FAD) | - Simple equipment - Nice structural contrast resulting from extracellular matrix on the tissue scale - Weak intracellular signals due to dependence on scattering intracellular fluorophores |
| Photoacoustic microscopy | Optical absorption and ultrasonic wave generation | - Nice matching with H&E staining on the tissue scale - Fast imaging - Insufficient spatial resolution for cellular imaging - Limited chemical information with dependance on molecules with strong optical absorption |
| Quantitative phase microscopy | Phase shift after light propagation | - Simple equipment - Lacking sharp intracellular contrast - High reliance on post-processing |
| Deep-ultraviolet microscopy | Optical absorption | - Simple equipment - High spatial resolution with the short wavelength of UV - Photodamage effect - Limited chemical information with dependance on molecules with strong ultraviolet light absorption |
| Stimulated Raman scattering microscopy | Molecular vibration modes | - Subcellular spatial resolution and clear distinguishment of subcellular structures (lipid droplets, nuclear membrane, etc.) - High chemical specificity and rich biomolecular information (lipid, protein, nucleic acid, etc.) - Requires specialized laser system |

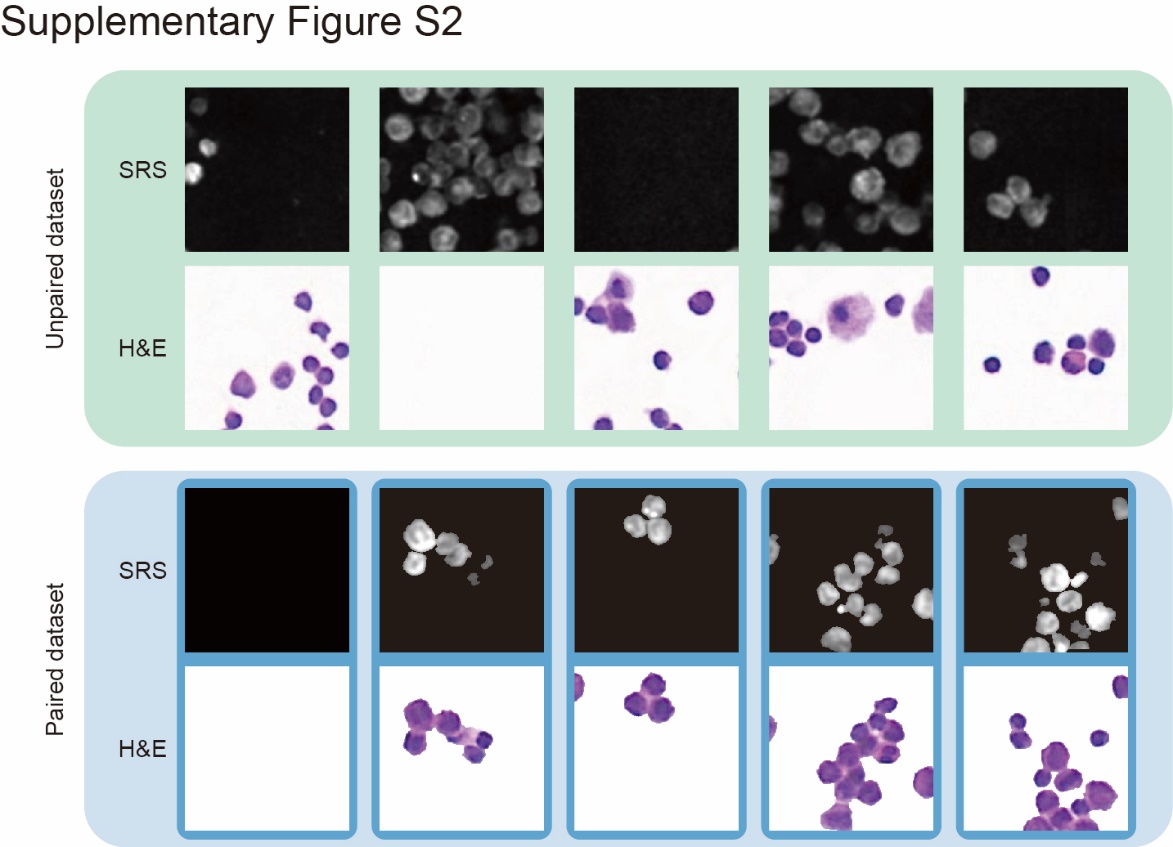

**Fig. S1 Diagram of unpaired and paired dataset.** Unpaired and paired dataset contained unregistered and registered patches of SRS and H&E-stained images, respectively. Slight remaining mismatches after registration were acceptable because the paired dataset was not used for model training.

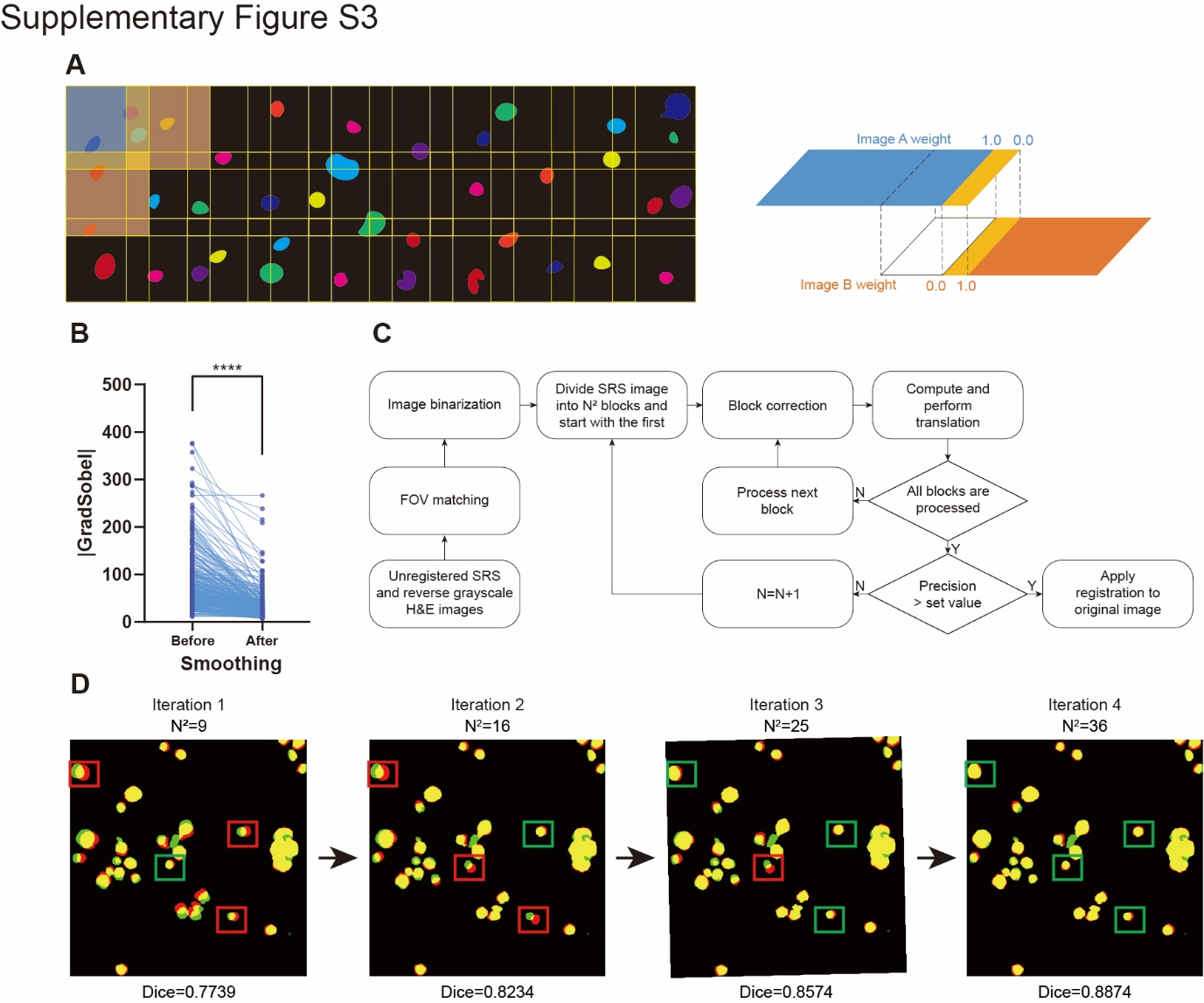

**Fig. S2 Principle and evaluation of image stitching and registration.** **A**, The schematic of image stitching. FOVs captured with a 30% overlap were stitched in the absolute Cartesian coordinate system. In one region with a certain width in the overlap, pixels were linearly blended, and either of the two adjacent FOVs determined the intensity in the other region in the overlap. **B**, Sobel gradients on the mosaic image seams before (mean±SD=67.20±71.58) and after smoothing (mean±SD=44.70±42.05), tested by Wilcoxon matched-pairs signed rank test (n=4815). Only pixels more intense than an empirical threshold were included to emphasize the smoothing effect. **C**, Image registration workflow. **D**, A representative process of iterative registration corresponding to steps in c after image binarization. Red area, the mismatched foreground of the SRS patch. Green area, the mismatched foreground of the H&E-stained patch. Yellow area, the matched foreground of two patches. ****: p<0.0001.

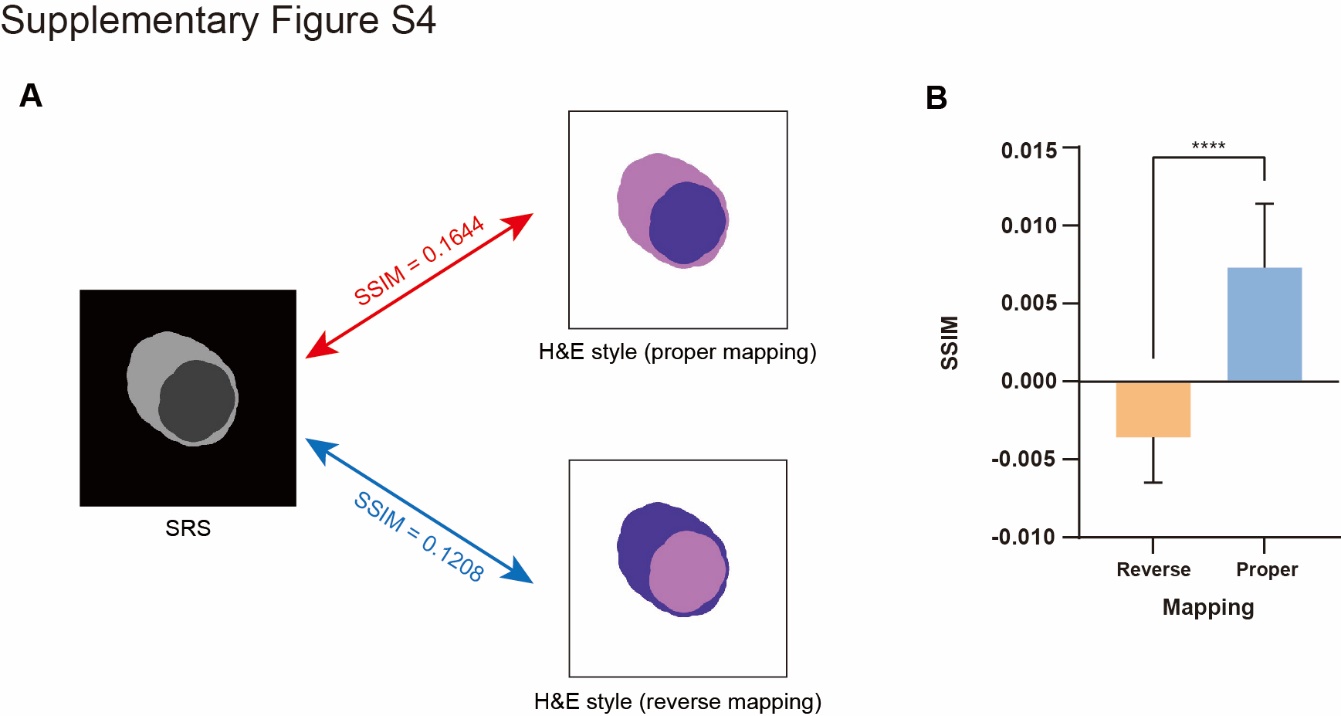

**Fig. S3 Hypothesis about the SSIM and its verification.** **A**. Comparison of SSIMs between a simulated SRS image and its corresponding simulated properly/reversely stained image. A hypothesis was proposed that properly-stained images were more structurally similar to input images than reversely stained ones. **B**, Verification of the hypothesis through representative reversely- (mean±SD=-0.0036±0.0029) and properly-stained (mean±SD=0.0073±0.0041) images produced by UTOM and CellGAN, respectively, tested by Wilcoxon matched-pairs signed rank test (n=147). ****:p < 0.0001.

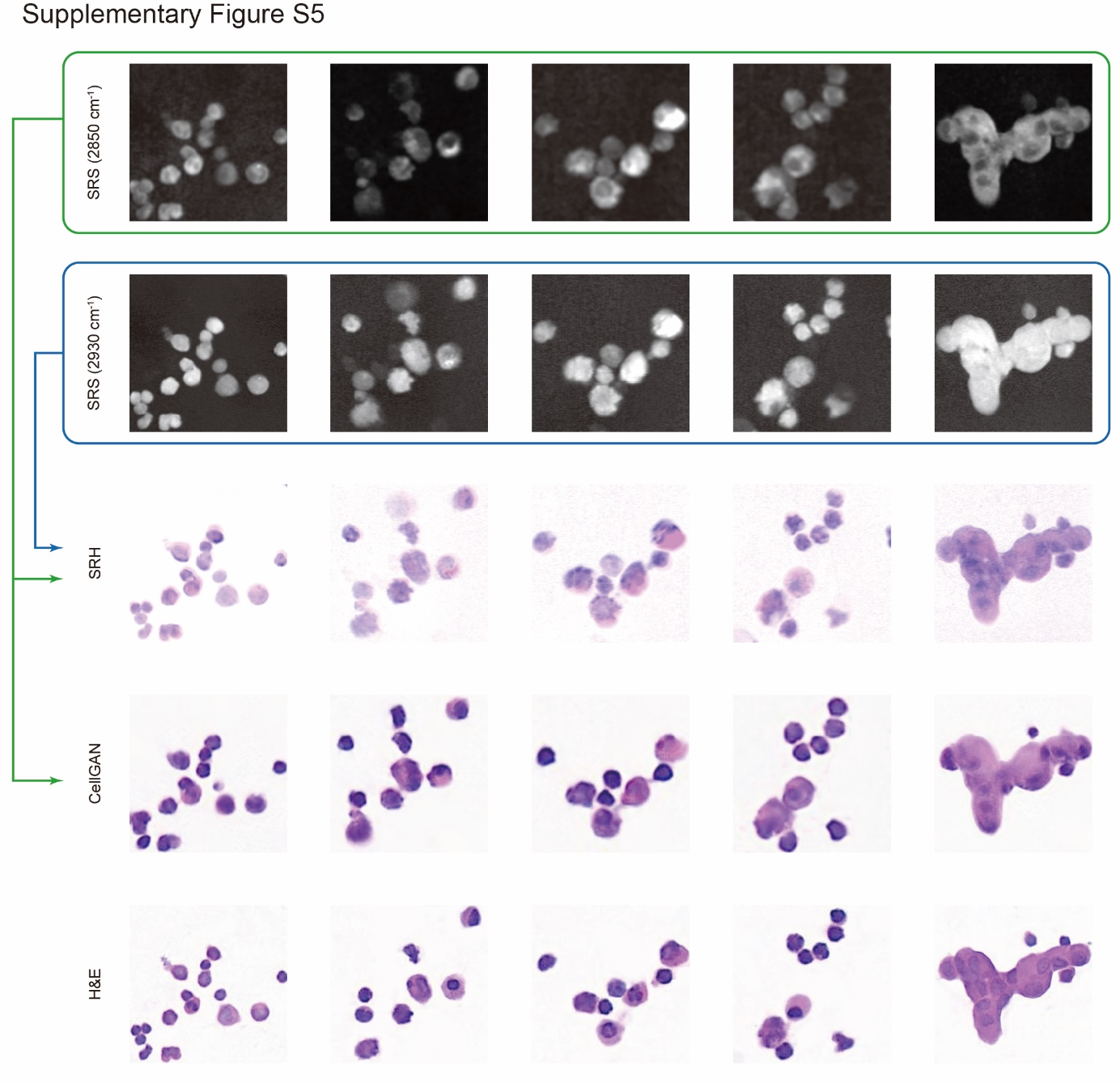

**Fig. S4 Comparison between CellGAN with single-color SRS and SRH with two-color SRS.** SRH images were calculated with SRS images at two Raman shifts, 2850 cm^-1^ and 2930 cm^-1^, in the same FOVs.

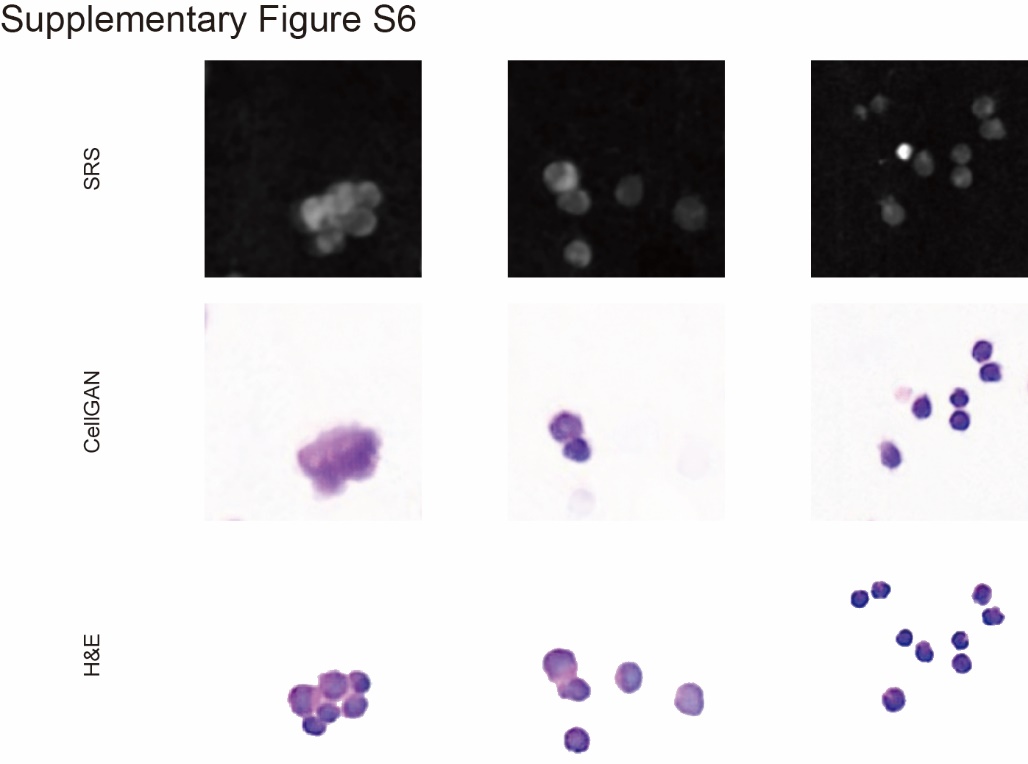

**Fig. S5 Artifacts in virtually stained cells.** Fusion of virtually stained cell cluster and neglected unclear cells caused by loss of focus are displayed.

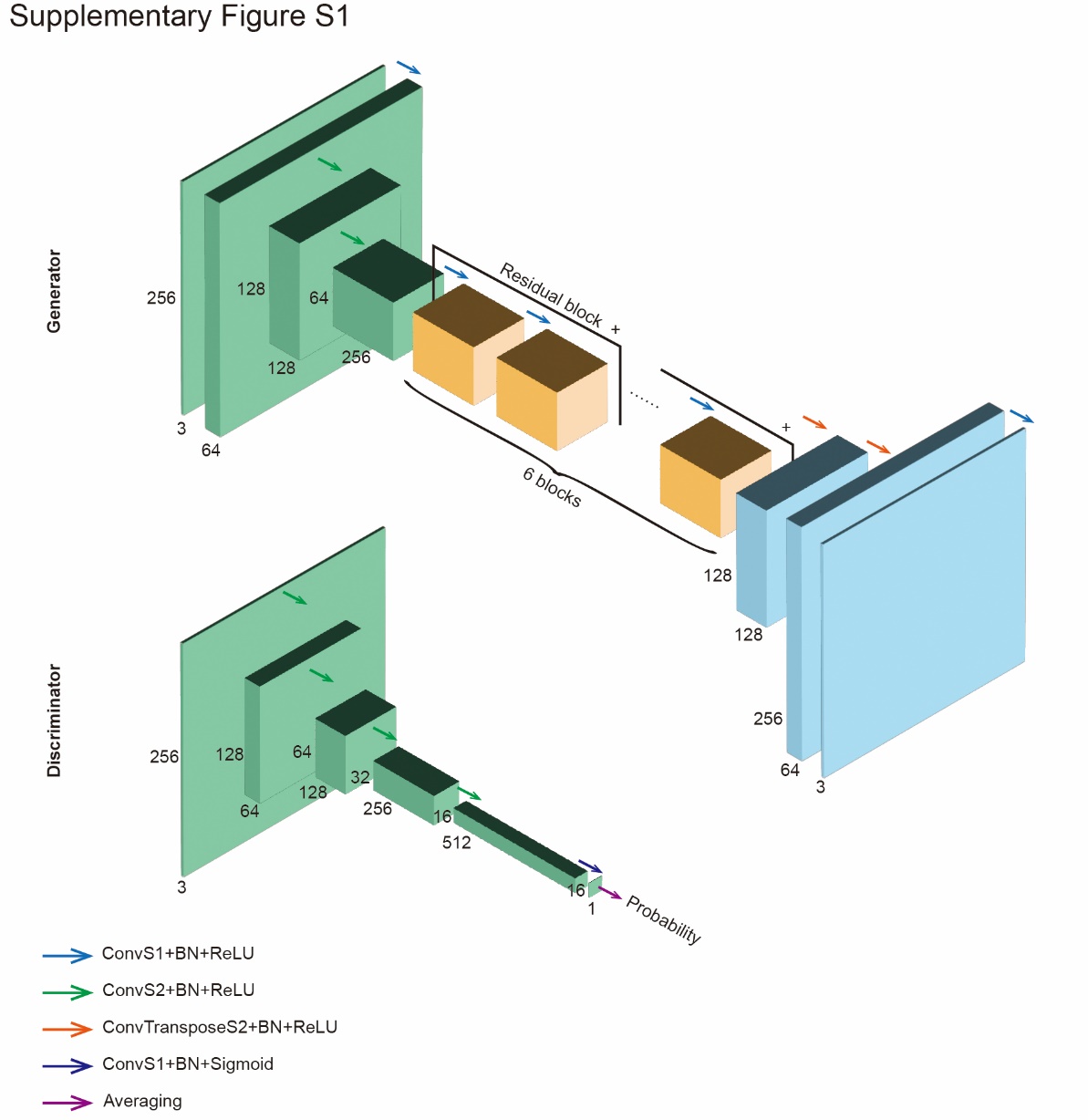

**Fig. S6 Architectures of generators and discriminators in CellGAN.** Generators were convolutional neural networks composed of encoders (green layers), residual blocks (orange layers), and decoders (cyan layers). Discriminators were PatchGANs outputting the average patch-wise prediction of an image, denoting the probability that the input image was collected from reality instead of synthesized by a generator. All inputs and outputs had 256×256 pixels and three channels.
